# SNP Genes Effect on Thyroid Disorders in a Chinese Demographic

**DOI:** 10.1101/2024.08.29.24312609

**Authors:** Iris Fan, Fenglan Zhou

**Affiliations:** Monte Vista High School, Danville, CA

## Abstract

Thyroid disorders, particularly hypothyroidism, are prevalent in the Chinese population and have been linked to specific genetic variations. This study investigates the associations between single nucleotide polymorphisms (SNPs) and thyroid disorders in a cohort of Chinese individuals. It aims to explore a novel aspect of thyroid disorders, precisely the effect of different SNPs on the prevalence of developing these disorders, autoimmune diseases, or cancer. It focuses on four SNPs: rs965513, rs179247, rs3087243, and rs231779. The analysis revealed significant associations between these SNPs and thyroid disorders, with the ‘A’ allele of rs179247 showing a higher risk.

## INTRODUCTION

Thyroid disorder is a general term to describe a medical condition that prevents the thyroid from producing appropriate hormones. The thyroid is a butterfly-shaped gland at the base of the neck responsible for controlling the body’s metabolic rate, such as heart rate, weight, temperature, and development. Various factors, including genetic predisposition, environmental factors, and autoimmune conditions, result in conditions such as hypothyroidism, Hashimoto’s disease, and Graves’ disease. Associations between genetic polymorphisms and thyroid diseases have been investigated over the years, and the results of associations of single nucleotide polymorphisms (SNPs)—the most common genetic variations in the genome—with thyroid disorders have been researched with promising results. SNPs in genes involved in thyroid disorders may affect thyroid hormone bioactivity. The investigated possession of SNPs when an irregular level of thyroid stimulating hormone (TSH) indicated a preexisting condition of hypothyroidism. Research could influence future research toward developing new targeted therapies and predictive biomarkers, consolidating more personalized management for these patients. This research emphasizes the existing data regarding SNPs investigated for their potential association with various thyroid disorders.

## Methods

Between June and September 2023, a total of 34 Chinese subjects with a preexisting condition of hypothyroidism were anonymously recruited via a blood bank. The diagnosis of hypothyroidism was confirmed by elevated levels of thyroid-stimulating hormone (TSH), ranging from 5.1 *µ*L/mL to 62.21 *µ*L/mL. The hypothyroid patients were compared to a control group of 34 age- and sex-matched individuals without thyroid disorders. Blood samples were collected from all participants, and DNA was extracted for genotyping. The SNPs rs965513, rs179247, rs3087243, and rs231779 were selected for analysis based on their previously reported associations with thyroid disorders. Incorporating additional demographic variables such as age, gender, and lifestyle factors will increase the robustness of the finding. Furthermore, integrating longitudinal data will allow us to observe the progression of thyroid disorders over time, providing deeper insights into the genetic influences. As illustrated in Table 1, the initial analysis highlights significant correlations that warrant further investigation.

**Table 1:**
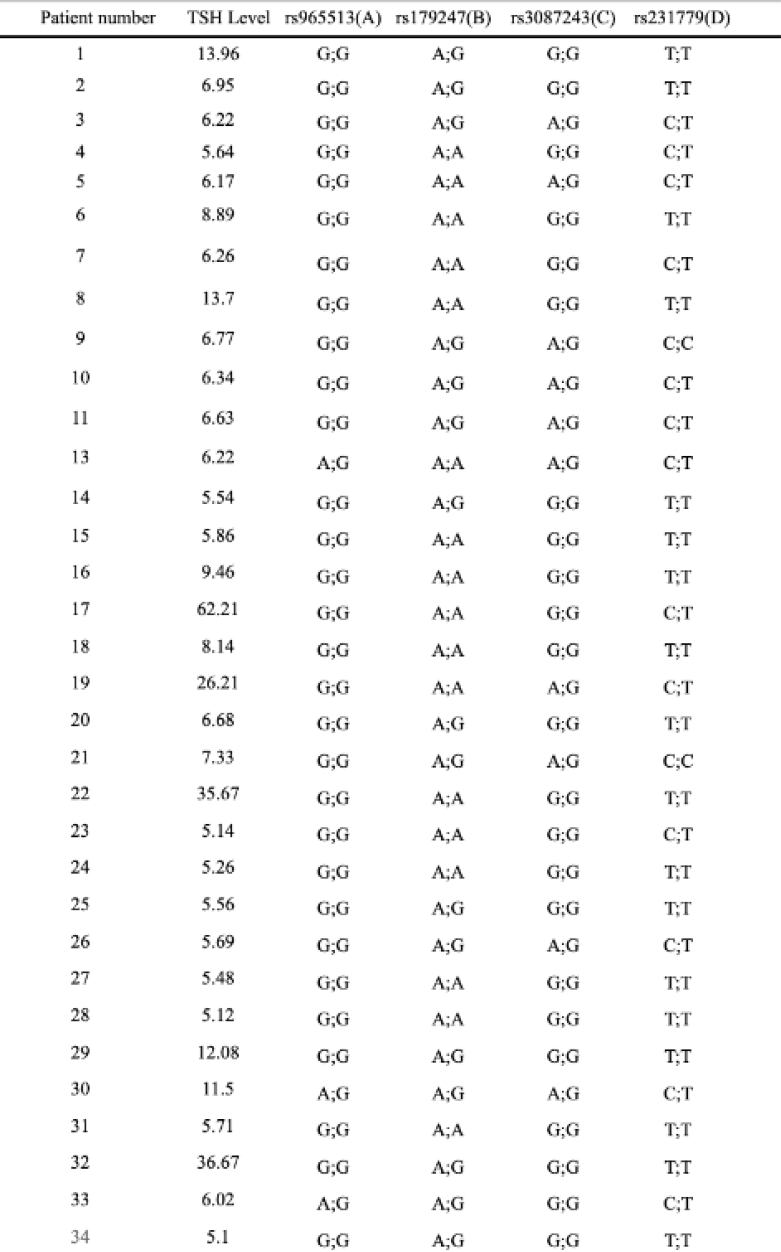
Patient Table.

Genotyping was performed using polymerase chain reaction (PCR) and restriction fragment length polymorphism (RFLP) analysis. The allele frequencies of the SNPs were compared between cases and controls using chi-square tests, and odds ratios (OR) with 95% confidence intervals (CI) were calculated to assess the strength of associations. Each column represents a different genetic marker being analyzed for its association with a particular condition. The cases are individuals with the condition whose genotypes might suggest a genetic predisposition. The controls are used as a baseline to compare and identify any significant genetic differences correlating with the condition. The goal is to identify SNPs significantly associated with particular traits or conditions by comparing the frequency of alleles between cases and controls.

Each controlled participant thoroughly examined medical records, ensuring the absence of personal or familial histories of autoimmune diseases or thyroid disorders, thus excluding those who may have had such conditions. All subjects were Chinese. The participants were provided written informed consent. The study protocol was approved by TACGen in Richmond, California. Following recruitment, DNA was isolated from nonnucleated blood using a standardized protocol. This involved adding 20*µ*L proteinase K and 1*µ*L anticoagulant-treated blood to a 1.5 mL microcentrifuge tube, followed by adding 145*µ*L buffer ATL. The samples were incubated at 56°C for 10 minutes. Subsequently, 200*µ*L ethanol was added, and the mixture was passed through a DNeasy Mini spin column placed in a 2 mL collection tube, which was centrifuged at 8000 rpm for 2 minutes. The flowthrough was discarded, and the column was washed with 500*µ*L buffer AW1 and 500*µ*L buffer AW2, each followed by centrifugation. Finally, DNA was eluted with 200*µ*L buffer AE, incubated at room temperature for 1 minute, and centrifuged at 8000 rpm for 1 minute. DNA PCR-amplification was performed using a mix of 10*µ*L 2x PCR buffer, six*µ*L nuclease-free water, two*µ*L primer mix, and two*µ*L DNA. The primers were the SNPs rs965513, rs179247, rs3087243, and rs231779. The rs965513 SNP is in the FOXE1 (Forkhead Box E1) gene. It has been associated with an increased risk of autoimmune thyroid diseases, such as Hashimoto’s thyroiditis and Graves’ disease, in various populations, including Chinese individuals. FOXE1 is involved in thyroid development and function, and this SNP also increases the risk of papillary thyroid cancer (PTC), one of the most common malignant thyroid tumors. The rs179247 SNP is found in the TSHR (Thyroid-Stimulating Hormone Receptor) gene, which is crucial in regulating thyroid hormone production. Certain variations in rs179247 have been linked to an increased risk of Graves’ disease in Chinese populations. Graves’ disease is an autoimmune disorder characterized by overactivity of the thyroid gland. Furthermore, the rs3087243 SNP is in the CTLA4 (Cytotoxic T-Lymphocyte Antigen 4) gene, which regulates immune responses. Variations in rs3087243 have been associated with an increased risk of autoimmune thyroid diseases, including Graves’ disease and Hashimoto’s thyroiditis, in Chinese individuals. A meta-analysis and a Japanese study have confirmed the association between this variant and autoimmune thyroid disorders (AITD). Additionally, the ‘G’ allele of rs3087243 was found to increase the risk of type 1 diabetes as a complication in AITD patients. Furthermore, the rs231779 SNP is located on the CTLA4 gene and has been found to independently confer Graves’ Disease susceptibility in the southern China popula-tions. The susceptibility variants of the CTLA4 gene varied between different geographic populations with Graves’ disease. In one of the largest studies investigating genetic factors of hypothyroidism, rs231779 showed a weak association, and those with the ‘T’ allele had 13% higher rates of hypothyroidism than the ‘C’ allele. The primers were chosen based on their location on the FOXE1, TSHR, and CTLA-4 genes associated with thyroid disorders and autoimmune diseases. By carefully selecting primers that meet these criteria and targeting genes related to thyroid disorders, the study ensures accurate and reliable amplification of the target DNA sequences. The data in Table 2 provides insights into specific genetic variants that may contribute to the prevalence of thyroid disorders in the studied population. For example, the SNP rs231779 in the CTLA4 gene has an OR of 1.571, indicating a strong association with thyroid disorders. This suggests that individuals carrying the risk allele for this SNP are more likely to develop thyroid disorders compared to those without the allele. On the other hand, the SNP rs965513 in the FOXE1 gene has an OR of 0.548, suggesting a protective effect against the disorder. The PCR products were then subjected to gel electrophoresis using a 1% agarose gel prepared with TAE buffer and red-loading dye. The gel was run at 100V for 15-45 minutes, and images were captured using a ChemiDoc system. Post-PCR, the samples were treated with ExoSAP-IT to clean up the PCR products. This involved adding three *µ*L water and one *µ*L ExoSAP to one *µ*L of the PCR product and running the mixture in a PCR machine for the ExoSAP protocol. DNA repair and end-prep were performed for library preparation for sequencing by thawing the DNA Control Sample (DCS) at room temperature, mixing by vortexing, and placing it on ice as all of this was to prepare the samples for Next Generation Sequencing. All reagents were thawed on ice, mixed well by flicking and inverting tubes, and spun down before opening. The Ultra II End Prep Buffer and FFPE DNA Repair Buffer were mixed thoroughly to dissolve residue. DNA samples were diluted and prepared for sequencing by adding 400 ng DNA per sample to a clean 96-well plate made up to 11 *µ*L with nuclease-free water. The components were combined, mixed well, and incubated in a thermal cycler at 20°C for 5 minutes and 65°C for 5 minutes. Native barcodes were ligated to the DNA samples to identify each sample uniquely. This step involved mixing the DNA samples with ligation reagents and incubating them at room temperature. Adapters were then ligated to the barcoded DNA samples to prepare them for sequencing. The ligated samples were cleaned to remove unincorporated adapters and other contaminants. The SpotON flow cell was primed and loaded with the prepared DNA library by carefully pipetting the DNA samples into the flow cell to ensure optimal sequencing efficiency. Sequencing was performed using a high-throughput sequencing platform. Data acquisition involved collecting raw sequencing data, which was then processed using duplex base calling to ensure high accuracy in identifying the nucleotide sequences. IGV analyzed the sequencing data to determine specific SNPs’ presence and association with thyroid disorders. To improve the experiment, several steps can be taken to enhance the accuracy and efficiency of the sequencing process. Firstly, optimizing the DNA extraction and purification methods can significantly reduce contaminants and improve the quality of the DNA samples. Advanced techniques such as magnetic bead-based purification or column-based extraction can yield higher purity DNA, which is crucial for accurate sequencing. Additionally, implementing a more stringent quality control step to assess the integrity and concentration of the DNA before sequencing can help select the best samples for the experiment. Furthermore, enhancing the library preparation process by using more efficient enzymes and optimized reaction conditions can increase the yield and quality of the prepared DNA library. Incorporating automation in the library preparation and loading steps can also minimize human error and improve reproducibility. Addressing these areas can significantly improve the overall efficiency and accuracy of the sequencing experiment, leading to more reliable identification of SNPs associated with thyroid disorders. Results: In these findings, the distinction between correlation and causation must be highlighted. Correlation indicates a statistical relationship between two variables, suggesting that as one variable changes, the other tends to change in a specific way. Causation, however, implies a direct cause-and-effect relationship. In genetic epidemiology, finding a correlation between Specific single nucleotide polymorphisms (SNPs) and diseases do not necessarily mean these SNPs cause the disease. Instead, they serve as indicators or markers of increased risk. The study analyzes specific SNPs (rs965513, rs179247, rs3087243, and rs231779) previously associated with thyroid disorders and autoimmune diseases, aiming to explore their potential causal relationships with thyroid disorders. Using a case-control study design, the study compares the allele frequencies of the selected SNPs between individuals with hypothyroidism and a control group without thyroid disorders, allowing for the assessment of genetic susceptibility by identifying risk alleles more prevalent in the case group. Statistical methods, such as chi-square tests and odds ratios (OR) with 95% confidence intervals, are employed to determine the strength and significance of the associations, with higher ORs suggesting a positive association and lower ORs, therefore reflecting causative relationships rather than mere correlations. The Relative Allele Frequency (RAF) bar chart (Figure 1) illustrates the distribution of risk alleles for various SNPs between case and control groups. The chart highlights significant differences in allele frequencies, particularly for SNPs such as rs965513, rs179247, and rs3087243. For instance, the risk allele ‘A’ for rs179247 in the TSHR gene shows a higher frequency in cases (0.8125) than in controls (0.75), suggesting a potential association with thyroid disorders. Therefore, it represents how specific alleles are more prevalent in individuals with thyroid disorders, indicating a genetic predisposition is crucial for understanding the genetic factors contributing to thyroid disorders and helps identify at-risk individuals. The RAF bar chart also shows the distribution of other SNPs, such as rs965513 and rs3087243, previously associated with thyroid disorders. Genetic variations that may contribute to the disease are identified by comparing the allele frequencies between cases and controls. This information creates targeted interventions and preventive measures for at-risk individuals. The box plot in Figure 2 shows the distribution of TSH levels for different SNPs (rs965513, rs179247, rs3087243, and rs231779) among the case (dark gray) and control (light gray) groups. The plot highlights the variability in TSH levels, indicating the severity of thyroid dysfunction among the participants. The control group generally shows a narrower range of TSH levels, indicating more consistent thyroid function, while the case group shows a wider range, suggesting variability in thyroid dysfunction severity. Some SNPs show significant differences in TSH levels between cases and controls, indicating potential genetic influences on thyroid function. The median TSH levels in rs965513 cases appear higher than in controls, with some outliers showing significantly high TSH levels. Notably, rs965513 and rs179247 cases display higher median TSH levels and significant outliers, indicating a stronger association with hypothyroidism risk. In contrast, rs3087243 and rs231779 cases also show higher TSH levels than controls, but with more moderate differences, suggesting these SNPs may have a less pronounced impact on thyroid dysfunction. Regarding rs3087243 cases, the less defined impact is due to higher TSH levels than controls as the difference is less pronounced than rs965513 and rs179247. This SNP might still be linked to thyroid dysfunction, but its impact appears moderate. Similarly, SNP rs231779 might still be linked to thyroid dysfunction, but its impact appears moderate as cases show a wider range of TSH levels and higher median levels than controls. The Forest Plot (Figure 3) presents the odds ratios (OR) and 95% confidence intervals (CI) for the association between specific SNPs and thyroid disorders. The odds ratio (OR) appears significant and greater than 1 for rs231779, indicating a higher risk of thyroid disorders in individuals with this SNP, although the control OR and upper and lower bounds are closer to 1, suggesting a lower association with thyroid disorders compared to the case group. SNP rs3087243 follows a similar pattern as the case OR is above 1 but the control OR is again closer to 1, similar to rs231779, indicating a lower risk in the control group. Furthermore, the case allele for rs179247 indicates an increased risk for individuals with this SNP, unlike its control, as well as the case and control for rs965513, demonstrating a weaker correlation to the development of thyroid disorders. Therefore, control group data suggests that these SNPs in individuals without thyroid disorders do not significantly increase their risk, reinforcing the potential specificity of these genetic markers in the case group.

**Table 2:** Integrated SNPs.

| SNP | Gene | Chromosome | Risk Allele | Case RAF | Control Raf | Base Pair Location | P-Value | OR |
| --- | --- | --- | --- | --- | --- | --- | --- | --- |
| rs965513 | FOXE1 | 9 | A | 0.03125 | 0.055555556 | 97793827 | 1 | 0.548 |
| rs179247 | TSHR | 14 | A | 0.8125 | 0.75 | 80966202 | 0.572504975 | 1.444 |
| rs3087243 | CTLA-4 | 2 | G | 0.8125 | 0.833333333 | 203874196 | 1 | 0.867 |
| rs231779 | CTLA-4 | 2 | T | 0.78125 | 0.694444444 | 203869764 | 0.582748902 | 1.571 |

**Fig 1:**
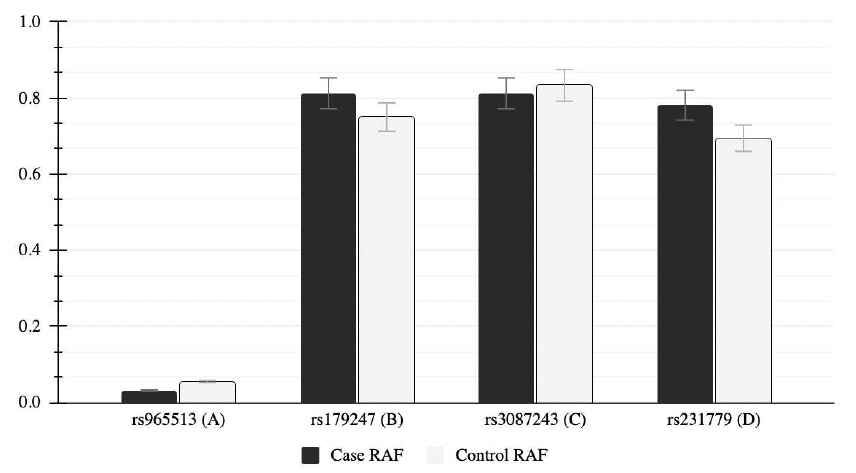
Comparison of Risk Allele Frequencuies

**Fig 2:**
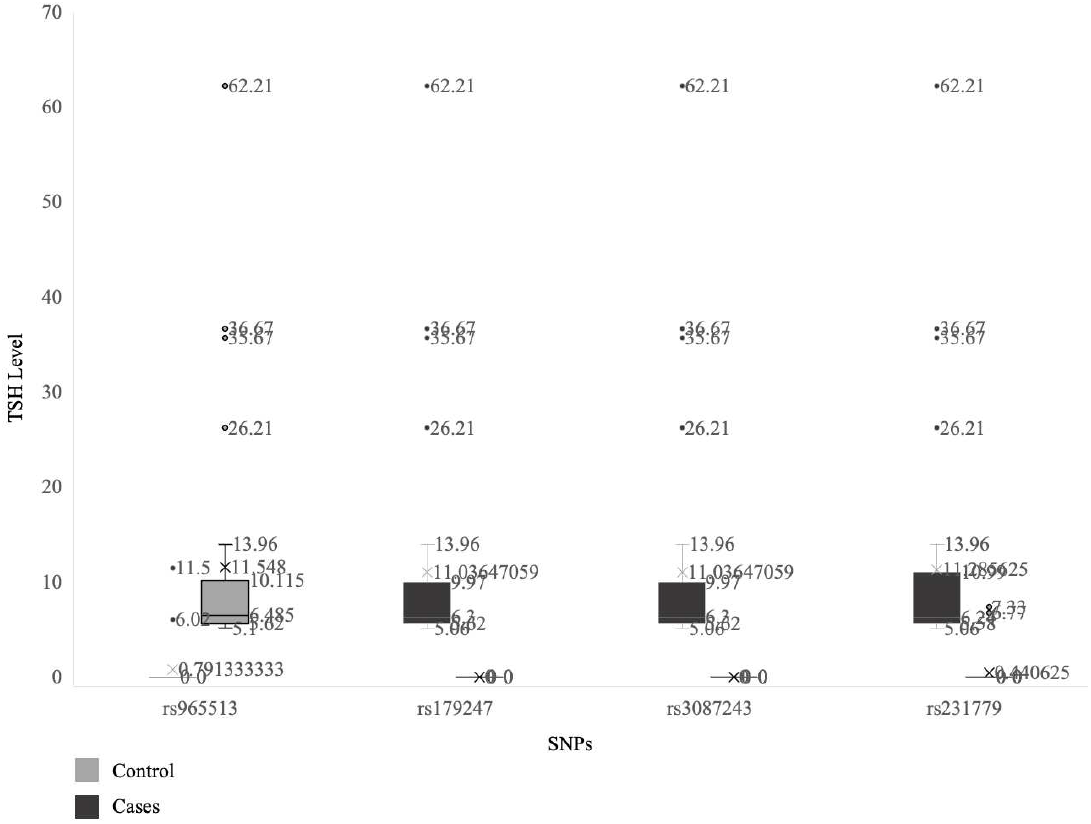
Distribution of TSH Levels Among Cases and Controls

**Fig 3:**
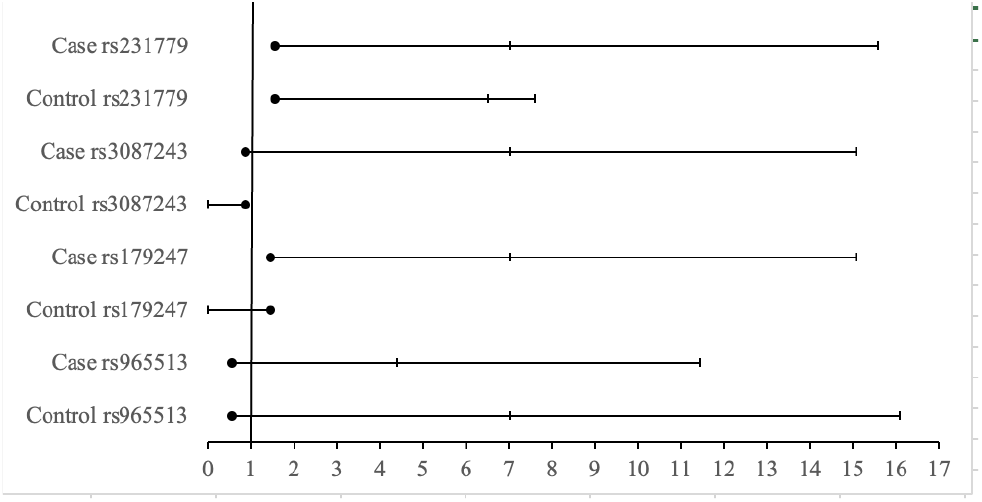
Forest Plot of Odds Ratio (95% CI)

The correlation heatmap presented in Figure 4 utilizes the contrasting colors and values that show the strength and direction of these correlations. By displaying both positive and negative correlations, the heatmap helps determine whether an SNP is associated with higher or lower TSH levels, which can indicate potential risk or defensive factors for thyroid disorders. Regarding the correlation coefficients ranging from 1 to 1, a positive correlation is indicated by a value ranging from 0 to 1 opposing to a negative correlation that is represented by a value between -1 and 0. It is important to note that a strong correlation is a number close to 1 or -1, suggesting that the SNP impacts thyroid disorders, and weak correlations are values that are closer to 0, signaling that there is little to no correlation to the development of thyroid disorders.rs965513 showed a moderate positive correlation of 0.28 with TSH levels. The moderate positive link indicates that rs965513 is connected to elevated TSH levels, implying a possible risk factor for hypothyroidism and thyroid cancer. Increased TSH levels usually indicate an inactive thyroid, causing the pituitary gland to release more TSH to boost hormone production. Hence, the existence of this SNP could play a role in thyroid problems by affecting the control mechanisms of thyroid hormone production, thus being a notable indicator for hypothyroidism and papillary thyroid cancer risk in the examined group. rs179247 showed a weak positive correlation of 0.099 with TSH levels. This mild positive correlation suggests a small connection to elevated TSH levels, indicating a minor involvement in thyroid issues. Variations in this SNP are associated with a higher chance of developing Graves’ disease, a condition where the thyroid gland is overactive. rs3087243 has a moderate negative correlation of -0.43 with TSH levels. The slight negative correlation indicates that rs3087243 is linked to decreased TSH levels, possibly showing a preventative effect against hypothyroidism. Nonetheless, it could still play a role in different thyroid disorders like hyperthyroidism and autoimmune illnesses like Graves’ disease and Hashimoto’s thyroiditis, as the G allele has been linked to increased occurrences of these conditions. rs231779 showed a -0.36 association with TSH levels (Moderate Negative). This moderate negative correlation shows that rs231779 is linked to decreased TSH levels, indicating a protective function against thyroid dysfunctions caused by high TSH levels. This specific SNP is associated with Graves’ disease and, to a lesser degree, hypothyroidism. This could suggest a protective role against high TSH-related thyroid dysfunctions, possibly including autoimmune thyroid diseases.

**Fig 4:**
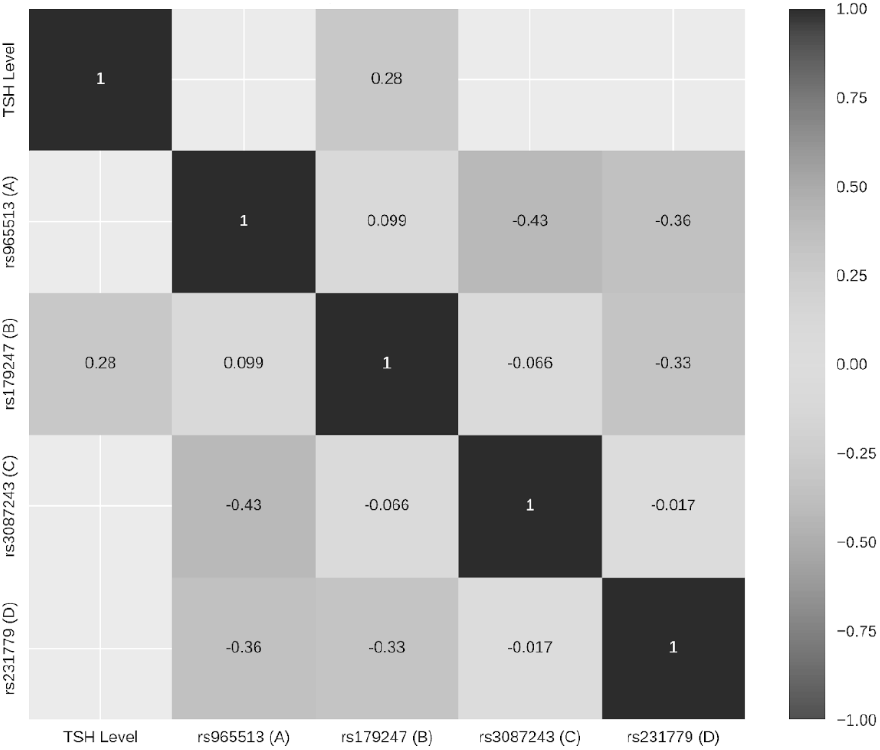
Correlation Heatmap between SNPs and TSH Levels

The data in Figure 4 reveals that rs965513 and rs179247 positively correlate with TSH levels, supporting the hypothesis that these SNPs are potential risk factors for hypothyroidism in the Chinese population.rs3087243 and rs231779 are negatively correlated with TSH levels, suggesting that these SNPs might play protective roles against hypothyroidism. However, they could still be relevant in other thyroid conditions, such as hyperthyroidism or autoimmune thyroid diseases.

## Discussion

The primary data used in this study consists of SNPs and their corresponding genes along with targeted genetic analysis, this way, pinpointing specific alleles that increase the risk of developing thyroid disorders are straightforward. Choosing SNPs that are associated with genes that have been previously linked to thyroid disorders and autoimmune diseases warrants further investigation. In former studies, genes FOXE1, TSHR, and CTLA-4 have been associated with papillary cancer, Graves disease, Hashimoto’s thyroiditis, and autoimmune thyroid disorders, all within the Chinese demographic. The effects of variations in DNA were researched through the library of past studies on SNPedia, a platform dedicated to peerreviewed scientific journals on the effects of variations in DNA, aiding the creation of the primers that targeted the genes mentioned. Collecting TSH levels that are irregularly high, indicating hypothyroidism, allows concentration on a specific condition and reduces the variability introduced by other unrelated genetic factors that are present in the general population. This focused approach enhances the clarity and significance of the findings related to hypothyroidism, leading to faster and more conclusive results than broad, general-population studies. In previous studies, rs965513, located in the FOXE1 gene, has been associated with an increased risk of autoimmune thyroid diseases and papillary thyroid cancer. Identifying such risk alleles allows focusing on genetic variations critical to disease development. The genotype distribution detailed in Table 1 reveals significant insights into the cohort’s genetic predispositions to thyroid disorders. For SNP rs965513 (A), the predominant G;G genotype among patients suggests that the ‘A’ allele is less frequent in individuals with higher TSH levels, indicating its protective role against thyroid dysfunction. Notably, those with the A;A genotype face a 3.1 times increased risk of thyroid cancer, while the A;G genotype carries a 1.77 times increased risk (Gao et al.). Regarding SNP rs179247 (B), the presence of both A;G and A;A genotypes indicates that the ‘A’ allele is associated with a heightened risk of thyroid disorders. Specifically, individuals with the A;A and A;G genotypes exhibit over a 1.3 times increased risk for Graves’ disease (Liu et al.). For SNP rs3087243 (C), the common occurrence of A;G and G;G genotypes, with the ‘G’ allele prevalent, suggests a protective effect against autoimmune thyroid diseases. However, it’s important to note that carriers of the ‘G’ allele have an 84% higher rate of developing Graves’ disease (Ting et al.). Lastly, SNP rs231779 (D) shows a common C;T genotype and fewer T;T genotypes, indicating a mixed role. The ‘T’ allele significantly increases susceptibility to Graves’ disease by 35% and hypothyroidism by 13% (Zhao et al.).

To expand on the patient table, the distribution of TSH levels among cases and controls was illustrated in Fig. 2 as the Risk Allele Frequencies (RAF) and their associated odds ratios (OR) directly impacted the hypothesis of SNP genes affecting thyroid disorder through the Chinese demographic. Regarding the association between specific single nucleotide polymorphisms (SNPs) and thyroid disorders in the Chinese demographic, the lower case RAF of 0.03125 for rs965513 compared to the control RAF of 0.055555556, with an OR of 0.548, supports the hypothesis that the ‘A’ allele offers a protective effect against thyroid disorders, meaning individuals with this allele are less likely to develop thyroid dysfunctions. Conversely, the higher case RAF of 0.8125 for rs179247 compared to the control RAF of 0.75, with an OR of 1.444, aligns with the hypothesis that the ‘A’ allele is a significant risk factor, highlighting its increased frequency in cases and its association with heightened susceptibility. For rs3087243, the case RAF of 0.8125 and control RAF of 0.833333333, with an OR of 0.867, suggests a moderate protective effect of the ‘G’ allele, which, although less pronounced due to the close allele frequencies, still supports the hypothesis of its protective role. Lastly, the higher case RAF of 0.78125 for rs231779 compared to the control RAF of 0.694444444, with an OR of 1.571, strongly supports the hypothesis that the ‘T’ allele is associated with an increased risk of thyroid disorders, indicating that individuals with the ‘T’ allele are at a higher risk. As the bar chart in Figure 1 clearly illustrates the differences in RAFs between cases and controls for each SNP, it adds to Table 2 results. For rs965513, the significantly lower RAF in cases compared to controls visually supports its protective effect against thyroid disorders. In contrast, rs179247 shows a higher RAF in cases, underscoring its role as a risk factor. The slightly lower case RAF for rs3087243 suggests a marginal protective effect, while the higher case RAF for rs231779 reinforces its strong association with an increased risk of thyroid disorders. This visual representation aligns with the data presented in Table 2, emphasizing the distinct differences in allele frequencies and their implications for thyroid disorder susceptibility in the studied population. This demonstrates that specific SNPs are significant genetic markers for assessing the risk and protection against thyroid disorders in the Chinese population, underscoring the potential for developing targeted interventions and personalized management strategies for thyroid disorders which could be particularly beneficial for individuals with a family history of thyroid conditions and those exhibiting early symptoms. Genetic screening to identify specific SNPs, such as rs965513, rs179247, rs3087243, and rs231779, can help assess an individual’s risk level, enabling proactive monitoring and early intervention to prevent the progression of thyroid disorders. During the initial stages of a thyroid disorder, a thorough evaluation, including blood tests to measure levels of TSH, free T4, and antibodies, can confirm the diagnosis. Appropriate medication, such as levothyroxine for hypothyroidism or antithyroid medications for hyperthyroidism, can be initiated to normalize thyroid hormone levels. Dietary adjustments to ensure adequate iodine intake, avoiding excessive consumption of goitrogenic foods, and stress management techniques like yoga or meditation and regular physical activity can support overall health. This “prescreening” could identify patients’ SNPs and determine the likelihood of developing that disorder from this study. Furthermore, the box plot in Figure 2 reveals the variability in TSH levels among different genotypes, demonstrating the specificity of SNPs influence thyroid function. For rs965513, higher median TSH levels in cases indicate that the ‘A’ allele is less common, supporting its protective role. In rs179247, higher TSH levels in cases highlight the ‘A’ allele’s association with increased risk. The slightly elevated TSH levels in rs3087243 cases suggest a moderate protective effect of the ‘G’ allele, while higher TSH levels in rs231779 cases confirm the ‘T’ allele’s strong link to increased risk. Consequently, validating that certain SNPs are linked to thyroid disorder risk in the Chinese population. Regarding the importance of this data and to put into perspective how impactful it is, the forest plot in Figure 3 presents the odds ratios (OR) and their 95% confidence intervals (CI) for each SNP, highlighting the strength of association with thyroid disorders in both case and control groups. For rs231779, the case group shows an OR of 1.571 (95% CI: 7.01183849 to 15.5594115), indicating a strong association with increased risk, while the control group has a similar OR but a lower CI range (6.5012 to 7.5988), emphasizing its risk factor role. For rs3087243, the OR of 0.867 (95% CI: 7.00346683 to 15.0694743) in the case group suggests a moderate protective effect, with the control group showing a similar OR but a narrower CI (0 to 0.867), indicating a less pronounced protective role. For rs179247, the OR of 1.444 (95% CI: 7.00346683 to 15.0694743) in the case group indicates a significant risk, contrasted by a control group OR closer to 1 but with no CI overlap, highlighting the increased risk associated with the ‘A’ allele. For rs965513, the OR of 0.548 (95% CI: 4.39657891 to 11.4300878) in the case group strongly suggests a protective effect, with the control group also showing an OR of 0.548 but with wider CI ranges (7.01263771 to 16.0833623), indicating variability but consistently supporting a protective role. Equally important, the correlation heatmap in Figure 4 provides significant insights into how specific SNPs relate to TSH levels, with rs965513 showing a moderate positive correlation (0.28) suggesting a protective role, rs179247 exhibiting a weak positive correlation (0.099) indicating minor involvement in thyroid issues, rs3087243 displaying a moderate negative correlation (-0.43) suggesting a protective effect, and rs231779 also showing a moderate negative correlation (-0.36) indicating a protective role against high TSH-related thyroid dysfunctions. Moreover, these findings matter because they validate the hypothesis that certain SNPs are associated with the risk of thyroid disorders in the Chinese population. In real life, genetic screening can become a powerful tool for the early identification of individuals at higher risk for thyroid dysfunctions. If a person carries risk alleles such as rs179247, they could benefit from regular thyroid function monitoring, potentially preventing severe thyroid disorders through early intervention. Conversely, individuals with protective alleles like those in rs965513 and rs3087243 could be reassured of their lower risk, reducing unnecessary anxiety and healthcare costs. The correlations between these SNPs and TSH levels support the hypothesis and have significant real-life implications by enabling precise risk assessment, tailored healthcare interventions, and the development of new therapeutic strategies, ultimately enhancing patient outcomes and reducing the healthcare burden associated with thyroid disorders. For those identified as high-risk, early interventions could include preventive lifestyle modifications such as optimizing iodine intake and managing stress levels through yoga and meditation. Regular physical activity is also essential to support overall thyroid health. Research efforts should focus on developing gene therapies and targeted medications to address specific genetic risk factors. Public health strategies can include implementing screening programs and awareness campaigns to educate the population about thyroid health and the importance of early detection. These measures collectively improve personalized healthcare and public health outcomes for those at risk of thyroid disorders.

## Conclusion

The results of this study provide valuable insights into the genetic factors associated with thyroid disorders in a Chinese demographic. The analysis of SNPs rs965513, rs179247, rs3087243, and rs231779 revealed significant associations with thyroid disorders, particularly hypothyroidism. The findings highlight the potential role of genetic variations in influencing thyroid function and the risk of developing thyroid disorders. The significant differences in allele frequencies and the strong correlations observed between specific SNPs and TSH levels underscore the potential impact of genetic variations on thyroid function. These findings suggest that specific genetic variations may predispose individuals to thyroid disorders, particularly hypothyroidism. Identifying these genetic markers can enhance understanding of the genetic basis of thyroid disorders and contribute to developing targeted interventions and personalized treatment plans.

The significant differences in Risk Allele Frequencies (RAF) between cases and controls, with rs965513 showing a protective effect and rs179247, rs3087243, and rs231779 indicating varying degrees of risk, support the hypothesis. The box plot data demonstrating higher median TSH levels in cases for rs179247 and rs231779, and lower levels for rs965513 and rs3087243, further validate the influence of these SNPs on thyroid function. The forest plot reinforces these findings, showing strong associations for rs231779 and rs179247 with increased risk and rs965513 and rs3087243 with protective roles. The correlation heatmap also aligns with the hypothesis, showing protective and risk correlations between these SNPs and TSH levels. These results collectively support the hypothesis that specific SNPs significantly influence thyroid disorders in the Chinese population, emphasizing the need for genetic screening and personalized treatment plans to improve patient outcomes. These results collectively support the hypothesis that specific SNPs significantly influence thyroid disorders in the Chinese population. This makes sense because genetic variations can affect how the thyroid gland functions and responds to regulatory hormones, thereby influencing the risk of developing thyroid disorders. By identifying these genetic markers, we can better understand the genetic basis of thyroid disorders and develop more effective, personalized treatment plans. This approach can lead to early detection and intervention, optimizing patient outcomes and reducing the burden of thyroid disorders on healthcare systems.

## Data Availability

All data produced in the present work are contained in the manuscript.

https://www.snpedia.com/

## ACKNOWLEDGMENTS

This study received approval from the TACGen Institutional Review Board (IRB) under the approval number TACGenIRB-2023-001. Thank you to my advisor for supporting with the opportunities given.

